# Within and between classroom transmission patterns of seasonal influenza among primary school students in Matsumoto city, Japan

**DOI:** 10.1101/2021.07.08.21259917

**Authors:** Akira Endo, Mitsuo Uchida, Naoki Hayashi, Yang Liu, Katherine E. Atkins, Adam J. Kucharski, Sebastian Funk

## Abstract

Schools play a central role in the transmission of many respiratory infections. Heterogeneous social contact patterns associated with the social structures of schools (i.e. classes/grades) are likely to influence the within-school transmission dynamics, but data-driven evidence on fine-scale transmission patterns between students has been limited. Using a mathematical model, we analysed a large-scale dataset of seasonal influenza outbreaks in Matsumoto city, Japan to infer social interactions within and between classes/grades from observed transmission patterns. While the relative contribution of within-class and within-grade transmissions to the reproduction number varied with the number of classes per grade, the overall within-school reproduction number, which determines the initial growth of cases and the risk of sustained transmission, was only minimally associated with class sizes and the number of classes per grade. This finding suggests that interventions that change the size and number of classes, e.g. splitting classes and staggered attendance, may have limited effect on the control of school outbreaks. We also found that vaccination and mask-wearing of students were associated with reduced susceptibility (vaccination and mask-wearing) and infectiousness (mask-wearing) and hand washing with increased susceptibility. Our results show how analysis of fine-grained transmission patterns between students can improve understanding of within-school disease dynamics and provide insights into the relative impact of different approaches to outbreak control.

**Significance:** Empirical evidence on detailed transmission patterns of influenza among students within and between classes and grades and how they are shaped by school population structure (e.g. class and school sizes) has been limited to date. We analysed a detailed dataset of seasonal influenza incidence in 29 primary schools in Japan and found that the reproduction number at school did not show any clear association with the size or the number of classes. Our findings suggest that the interventions that only focus on reducing the number of students in class at any moment in time (e.g. reduced class sizes and staggered attendance) may not be as effective as measures that aim to reduce within-class risk (e.g. mask-wearing and vaccines).

## Background

Influenza virus and other directly transmitted pathogens typically spread over social contact networks involving frequent conversational or physical contacts (1–4). There is evidence that schools are important social environments that can facilitate the transmission of influenza via close contacts between students (5–9). Previous studies have collected contact data between students using questionnaires and wearable sensor devices and found strong assortativity of contact rates within classes and grades (10–14), which is likely relevant to the within-school transmission dynamics of respiratory infections and the effectiveness of school-based interventions. However, such insights from contact data also need to be validated with real-world outbreak data because contacts as measured in those studies may not necessarily be fully representative of the types of contacts that lead to transmission (e.g. with regards to proximity and duration). In this light, the differential transmission rates of influenza associated with classes and grades have also been estimated from empirical outbreak data in a few studies (6, 15, 16). However, those studies are limited to the analysis of only one or two schools and included a relatively small number of cases (< 300). Therefore, robust findings across schools with different structures that capture the full range of heterogeneity in within-school transmission dynamics have remained a crucial knowledge gap.

Understanding how school population structures (e.g. class and school sizes) shape transmission dynamics is key to making predictions about outbreak dynamics and interventions in these settings. Modelling studies of school outbreaks often require a choice between the ‘density-dependent mixing’ and ‘frequency-dependent mixing’ assumptions (17). The density-dependent mixing assumes that the transmission rate between a pair of students is constant regardless of the class/school sizes, while the frequency-dependent mixing assumes an inverse proportionality between them. As a result, the reproduction number is expected to increase with class/school size with the density-dependent mixing assumption and remain stable with the frequency-dependent mixing assumption. Whether the transmission is best characterised by the density-dependent mixing, frequency-dependent mixing or any other alternative assumption may vary between different modes of transmission and exposure settings (18–22). However, choices between the assumptions made by existing studies of school outbreaks vary widely and are not based on a clear empirical concensus (9, 23–26). These makes it challenging to interpret simulation studies evaluating school-based interventions (e.g. reduced class sizes) because the estimated effect sizes can heavily rely on the assumed mixing patterns (27–31).

To fill this knowledge gap in heterogeneous transmission dynamics at school, we applied a mathematical model of influenza virus transmission to a large-scale dataset from the 2014-15 season in Matsumoto city, Japan, which included diagnosed influenza reports among 10,923 primary school students and their household members. The model accounted for within-school transmissions as well as introductions to and from households and risk from the general community, which constitute key social layers of transmission (32–34). Using this model, we estimated fine-scale heterogeneous transmission patterns among students within and between classes and grades, as well as determinants of transmission rates including school structures and precautionary measures.

## Results

We analysed citywide survey data of 10,923 primary school students (5–12 years old) in Matsumoto city, Japan in 2014/15, which included 2,548 diagnosed influenza episodes among students (Figure 1A). The dataset was obtained from 29 schools with a range of class structures (sizes and the number of classes per grade), allowing for detailed analysis of within and between class transmission patterns (Figure 1B). The attack ratio (i.e. the cumulative proportion diseased) in each school (excluding three distinctively small schools with fewer than 15 students per class) showed weak to null negative correlations with the mean class size and the mean number of classes per grade (Figure 1C). The onset dates of students showed a temporal clustering pattern associated with school structure (Figure 1D). When the students were partitioned into different levels of groupings (i.e. by class, grade, school and overall), the deviation of onset dates from the within-group mean tended to be smaller with finer groupings.

**Figure 1.**
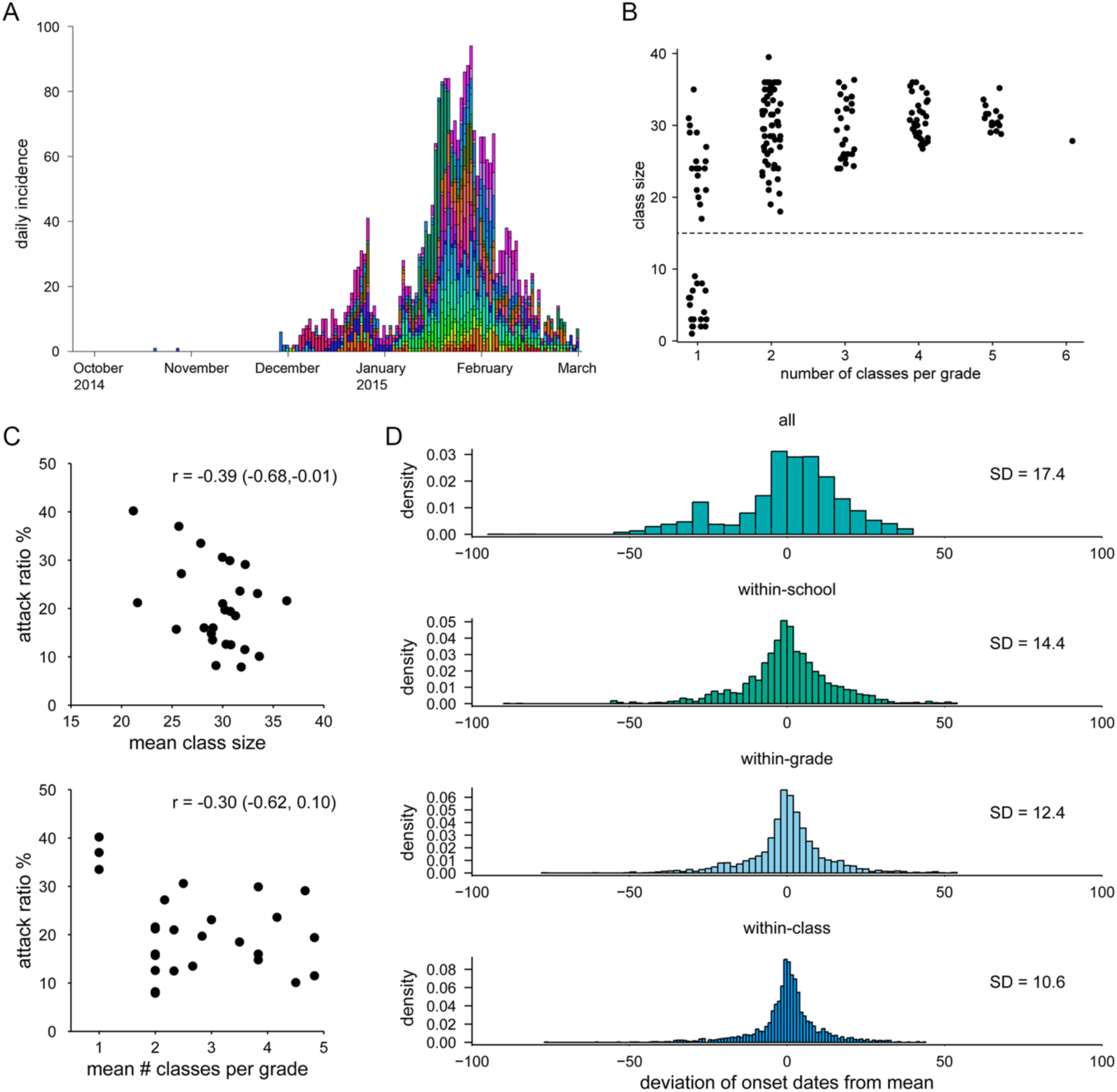
Transmission dynamics of seasonal influenza in primary schools in Matsumoto city, Japan and estimated effects of interventions for SARS-CoV-2. (A) Epidemic curve of seasonal influenza by illness onset in primary schools in Matsumoto city, 2014/15. Colours represent different schools. Month names denote the 1^st^ day of the month. (B) Scatterplot of the class sizes and the number of classes per grade in the dataset. Each dot represents a class in the dataset. Dots are jittered along the x-axis. Three schools had classes of fewer than 15 students (denoted by dotted horizontal line) and were excluded from the model fitting. (C) The scatterplots of the school attack ratio (%) against the mean class size and the mean number of classes per grade. The correlation indices (*r*) and the 95% confidence intervals are also shown. (D) Temporal clustering patterns of students’ onset dates with different levels of groupings reproduced from the school transmission model. The distributions of the deviance of each student’s onset from the group mean are displayed at overall, school, grade and class levels. The standard deviation (SD) of each distribution is also shown.

The temporal clustering shown in Figure 1D supports the hypothesis that the transmission is more likely within-class, followed by within-grade and within-school. We explored this further by estimating reproduction numbers within school. Using a mathematical model that accounts for different levels of interaction within and between classrooms and grades as well as introductions from households and community, we estimated the within-school effective reproduction number *R*_S_ of seasonal influenza in primary schools along with the breakdown of transmission risks associated with class/grade relationships (Figure 2). The relationship between any pair of students in the same school was classified as either “classmates”, “grademates” (in the same grade but not classmates) or “schoolmates” (not in the same grade). The estimated *R*_S_ was broken down as a sum of the contributions from these students, where the class size (*n*) and the number of classes per grade (*m*) were assumed to affect the risk of transmission. The reconstructed overall *R*_S_ in a 6-year primary school was estimated to be around 0.7–0.9 and was not significantly associated with *n* or *m* (Figure 2A). Namely, an infected student was suggested to generate a similar number of secondary cases irrespective of the class structure; although our estimates of *R*_S_ were about 15% smaller for the class size of 40 than 20^1^, the posterior p-value did not suggest a statistical significance (*p* ∼ 0.15 or above). As *R*_S_ was likely below 1 across class structures, school outbreaks may not have been sustained without continuous introductions from households and community. Transmission to classmates accounted for about two-thirds of *R*_S_ when each grade has only one class and was partially replaced by transmission to grademates as the number of classes per grade increases, while the sum of within-grade transmission (i.e. transmission to either classmates or grademates) remained stable (Figures 2B and 2C). Around 20–30% of overall *R*_S_ was explained by transmission to schoolmates throughout. We also obtained qualitatively similar results throughout our sensitivity analysis (Figure S3). In a 6-year school with 3 classes of 30 students, the risk of transmission was estimated to be 1.8% (95% credible interval (CrI): 1.4–2.5) from a given infected classmate of the same sex, 1.6% (1.2–2.1) the opposite sex, 0.13% (0.08–0.19) from a given infected grademate and 0.040% (0.029–0.055) from a given infected schoolmate (Table S2). The cumulative risk of infection from the community was estimated to be 2.2% (1.7–2.7) over the season.

**Figure 2.**
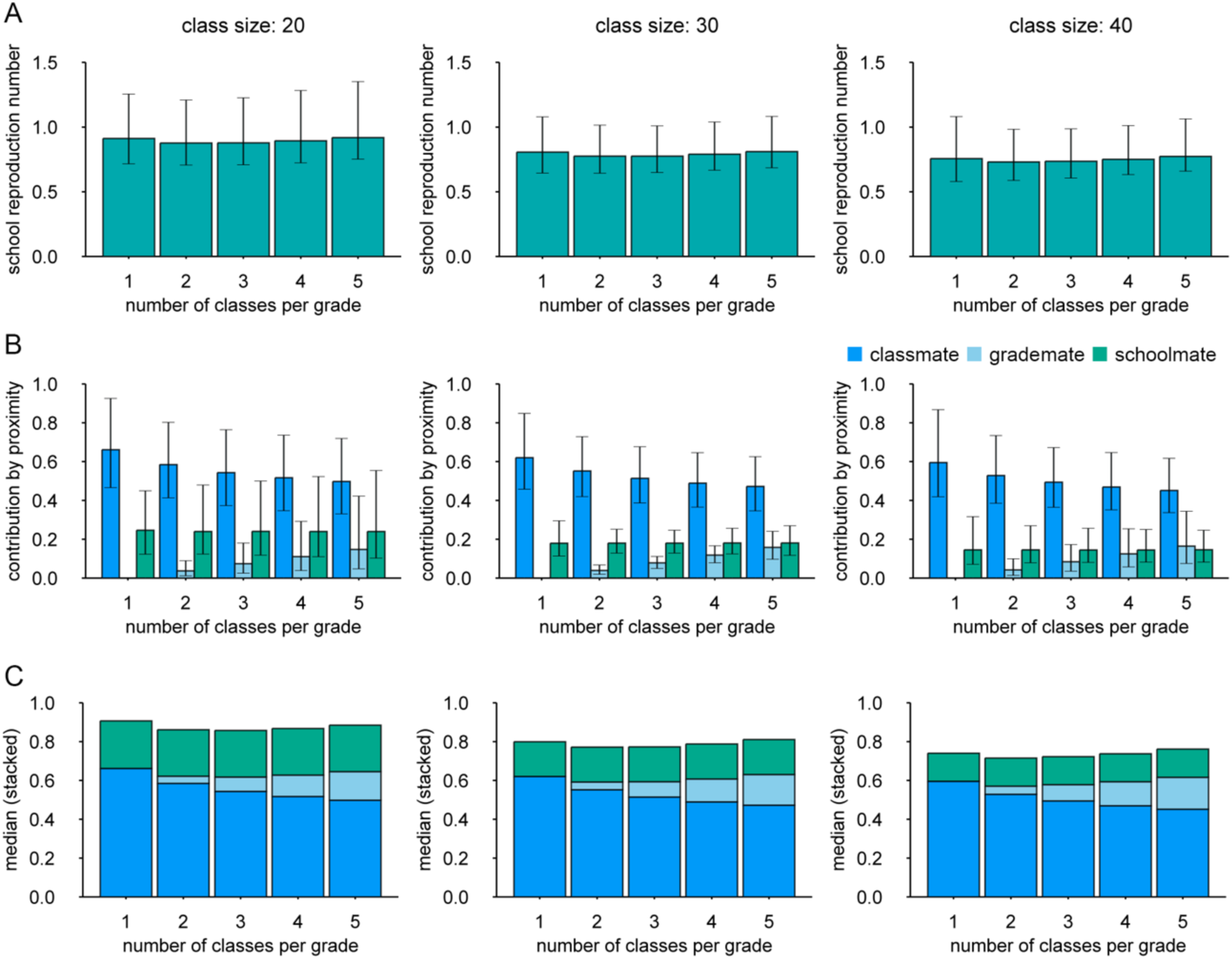
The estimated within-school transmission patterns of seasonal influenza among primary school students in Matsumoto city, Japan. (A) The overall school reproduction number (*R*_S_) under different class structures. Whiskers represent the 95% credible intervals (B) The breakdown of *R*_S_ corresponding to each type of within-school relationships. Whiskers represent the 95% credible intervals. Bottom panels: stacked graph of *R*_S_ based on the median estimates.

We incorporated a log-linear regression (35) into the above estimation of *R*_S_ to account for covariates that may affect the susceptibility or infectiousness of students. The results suggested that vaccines were associated with reduced susceptibility while mask wearing was associated with both reduced susceptibility and infectiousness (Table 1). Conversely, hand washing was associated with increased susceptibility. Reduced chance of transmission during the winter break (27 December 2014–7 January 2015) was captured as a 75% estimated decline in the infectiousness of cases whose onset dates were during the break. School grade, which serves as a proxy of students’ age, did not show a significant association with either susceptibility (relative value 1.10; CrI: 0.88–1.38) or infectiousness (relative value 0.78; CrI: 0.59–1.04).

**Table 1.** Covariates and effects estimated in the log-linear regression

| Covariate | Frequency in data | Relative susceptibility | Relative infectiousness |
| --- | --- | --- | --- |
| School grade (1 year increase) | — | 1.10 (0.88–1.38) | 0.78 (0.59–1.04) |
| Vaccine | 47.7% | 0.88* (0.81–0.96) | 0.97 (0.82–1.18) |
| Mask wearing | 51.4% | 0.77* (0.71–0.85) | 0.65* (0.55–0.78) |
| Hand washing | 80.1% | 1.54* (1.36–1.76) | 1.25 (0.96–1.65) |
| Onset in winter break | 5.9% (of cases) | — | 0.25* (0.15–0.39) |

Values are median estimates and 95% credible intervals.
\* Estimates with 95% credible intervals not crossing 1.

We estimated the breakdown of the source of infection for student cases based on the conditional probability predicted by the model and parameter estimates. The epidemic curve stratified by the estimated source of infection suggested that within-school transmission accounted for the majority of student cases while schools were open and that the within-household transmission was responsible for most of the cases reported during the winter break and shortly after (Figure 3A). The aggregated relative contribution suggested that 54.6% (CrI: 53.5–55.7), 38.7% (CrI: 37.9–39.5) and 6.7% (CrI: 5.9–7.5) of the student cases were acquired from school, household and community, respectively (Figure 3B).We estimated the possible relative effects of interventions altering the school population structure on the school reproduction number *R*_S_. We assumed that the estimated relative contributions of class/grade relationship to the transmission risk reflect the contact patterns between students which may also be relevant to the dynamics of another influenza outbreak at school (and potentially those of directly-transmitted disease outbreaks in general) and that the responses to interventions can be captured by the estimated relationship between *R*_S_ and the changes in the variables *n* and *m* according to each intervention (Table 2). Specifically, in the ‘split class’ scenario, each class was assumed to be split in half and taught simultaneously in separate classrooms, while in the ‘staggered attendance’ scenarios only half of the students attend school at the same time by introducing two different time schedules, e.g. morning and evening classes. The estimated relative effects of school-based interventions on *R*_S_ in a hypothetical setting of 6-year school with 2 classes per grade (40 students each) showed that splitting classes or staggered attendance alone was unlikely to reduce *R*_S_ (or may even be counteractive) (Figure 1D), which is consistent with the aforementioned estimates of *R*_S_ minimally associated with class sizes and the number of classes. By reducing interactions between students from different classes (so-called ‘bubbling’ or ‘cohorting’) by 90%, *R*_S_ could be reduced by up to around 20%. Combining split classes/staggered attendance with reduced interactions outside classes did not suggest incremental benefit in reducing *R*_S_. Given that these interventions typically require additional resources including staff and classrooms, the overall benefit to changing class structures for influenza control may be limited.

**Table 2.** Summary of interventions that changes the size/number of classes

| Interventions | Class size<br>($n$) | # classes per<br>grade ($m$) | Assumption |
| --- | --- | --- | --- |
| Baseline (‘no change’) | 40 | 2 | Students contact within and between classes and grades proportionally to the estimated transmission patterns in Figure 2. |
| Split class | 20 | 4 | Each class is split into two and taught simultaneously in separate classrooms. Students may contact each other between classes. |
| Staggered attendance<br>(within class) | 20 | 2 | Each class is split into two and taught separately in two different time slots (e.g. morning and evening). Students in different time slots do not contact each other and thus $R_S$ is calculated for students in one slot. |
| Staggered attendance<br>(between class) | 40 | 1 | Each class is allocated (as a whole) to either of the two different time slots and taught separately. Students in different time slots do not contact each other and thus $R_S$ is calculated for students in one slot. |

**Figure 3.**
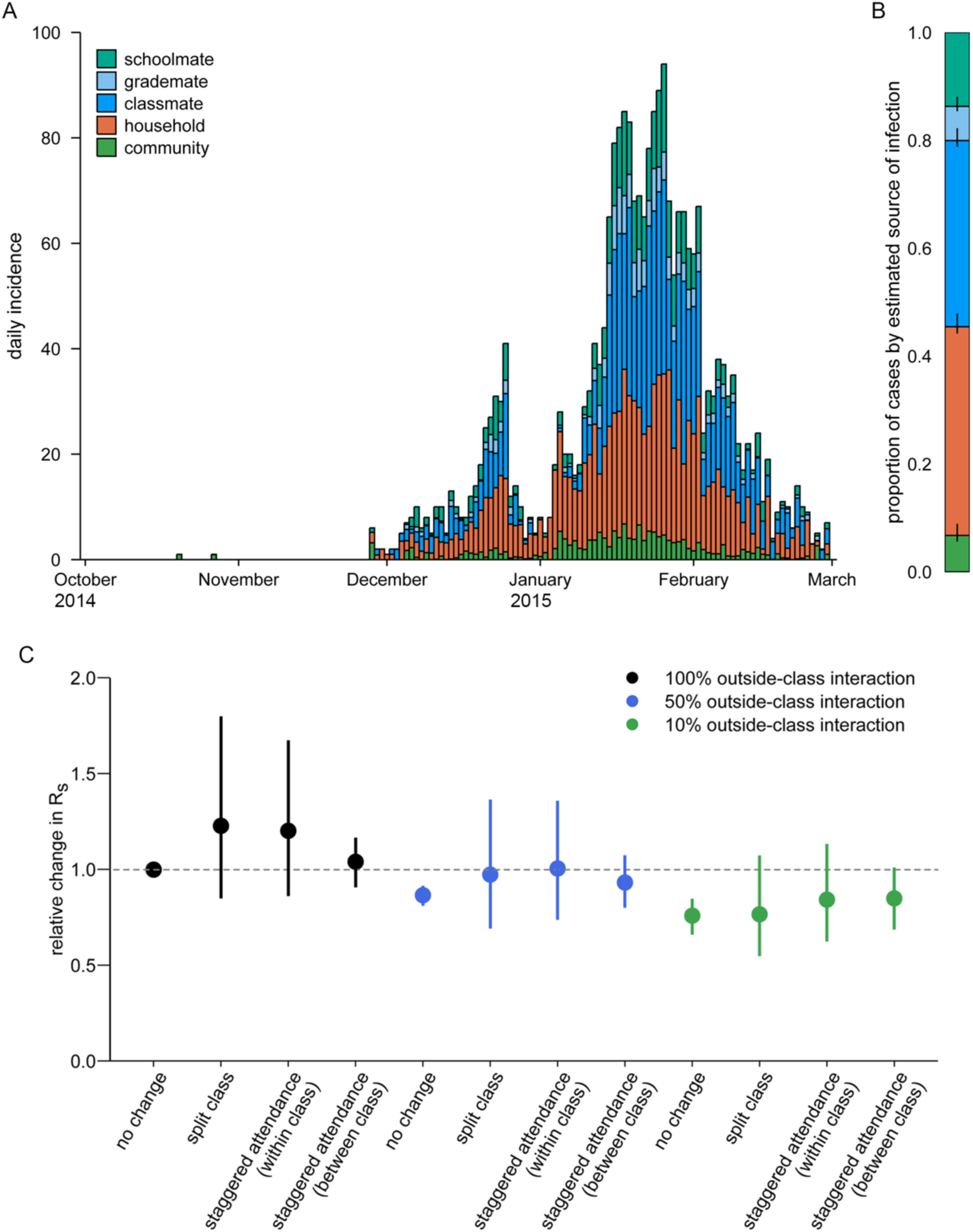
Reconstruction of students’ source of infection. (A) Epidemic curve stratified by the reconstructed source of infection. The conditional probability of infection from different sources was computed for each student and aggregated by date of illness onset. (B) Breakdown of the reconstructed source of infection. For each student, the source of infection was sampled based on the conditional probability to provide the proportion of students infected from each source. Bars denote posterior median and whiskers 95% credible intervals. (C) Expected relative changes in the school reproduction number under school-based interventions changing the structure of classes. Dots represent medians and whiskers 95% credible intervals. Reduced outside-class transmissions (i.e. from grademates or schoolmates) were also considered (50% reduction: blue; 90% reduction: green).

## Discussion

We used a mathematical model that stratified transmission within and between classes/grades to understand the dynamics of influenza transmission among primary school students. The inferred transmission dynamics of seasonal influenza in Matsumoto city, Japan, 2014-15 season suggested that the within-school reproduction number *R*_S_ stayed relatively constant regardless of the size or the number of classes (suggesting ‘frequency-dependent mixing’ (36)), in contrast to common modelling assumptions. The estimated *R*_S_ of 0.8–0.9, more than half of which was attributable to within-class transmissions, is consistent with a previous study in the United States (15). This value is also in line with the reported *R*_0_ of 1.2–1.3 for seasonal influenza (37) because our previous study estimated that the students in this dataset had infected 0.3–0.4 household members on average during this 2014-15 season (note that *R*_0_ corresponds to the overall number of secondary transmissions per student, including at school and household) (18). The value of *R*_S_ below 1 suggests that an outbreak cannot sustain itself within a school alone and that interactions through importing and exporting infections between households and the general community is likely to play a crucial role in the overall transmission dynamics. We estimated that school, household and community accounted for 55%, 39% and 7% of the source of infection for student cases, respectively. The attributable proportion was lower for schools and higher for households than the previous study (15), which may be explained by different scales of outbreaks in schools and households. In the Matsumoto city dataset, the overall attack ratio at school was lower (19%), students had larger households (average size 5.5) and there were more household cases than student cases (3996 vs 2548), as opposed to 35%, size of 3.4 and 141 vs 129 cases in (15).

The estimated breakdown of *R*_S_ revealed a number of notable patterns. As the number of classes per grade increased, the contribution of within-class transmission risk declined and was replaced by within-grade transmission. Combined with the almost constant overall *R*_S_, this might indicate that contact behaviour between students that contributed to transmission was only minimally affected by the student population density. That is, students may have had a certain number of ‘close friends’ with whom they had more intimate interactions that could facilitate transmission. In a school with more classes per grade, some of such friendship may have come from grademates instead of classmates, but the total number of close friends may have remained similar. This interpretation is in line with published evidence of influenza spreading predominantly in close proximity (38) and is likely to influence the expected effect of interventions not only for influenza but also other respiratory infectious diseases including COVID-19, which share similar routes and range of transmission (39, 40). Further disease-specific studies could elucidate the generalisability of these associations in more detail.

Our results suggested that interventions such as reducing class sizes or the number of students present (staggered attendance) may not be effective in constrast to what would be expected under the density-dependent mixing assumption (27–31). If interventions altering class structures are not accompanied by additional precaution measures and students try to resume their ‘natural’ behaviours (i.e. the same contact patterns as those in school with the resulting class structures) through so-called social contact ‘rewiring’ (41), the effect of such interventions can diminish or even reverse. For example, if other classes are absent due to staggered attendance, students may increase their interactions with classmates instead of their previous close friends in other classes. Our results are also consistent with a recent study of interventions against COVID-19 in US schools that did not find a significant risk reduction associated with reducing class sizes (42). Given the additional logistical resources required to implement these interventions, we propose that reducing the class sizes or the number of attending students should be considered only if they enable effective implementation of precaution measures such as physical distancing, environmental cleaning or forming social bubbles.

Using a log-linear regression analysis combined with a transmission model, we identified several precautionary measures associated with the susceptibility or infectiousness of students. Vaccines were associated with reduced susceptibility and masks with a reduction in both susceptibility and infectiousness. Influenza vaccine effectiveness in the 2014-15 season was suggested to be particularly low in Japan due to vaccine mismatch and estimated to be 26% (95% CrI: 7–41%) for primary-school-age children (6–12 years old) (43). Our estimate of a relative susceptibility of 0.88 (CrI: 0.81–0.96) in vaccinated students, which translates into a vaccine effectiveness of 12% (CrI: 9– 19%), is broadly consistent with this prior estimate. While existing evidence for the effectiveness of mask policies for the control of respiratory infections is still limited (44, 45), our estimates of small protective effects acting on the relative susceptibility (0.77; CrI: 0.71–0.85) and infectiousness (0.65; CrI: 0.55–0.78) lie within a plausible range based on evidence available to date (44, 46–48). Increased susceptibility associated with hand washing in our analysis, however, does not align with existing findings (49, 50). Although the underlying cause for this is unclear, the original report on the Matsumoto city dataset also reported a higher odds ratio (1.4; CrI: 1.27–1.64; unadjusted for differential exposure) and attributed it to the possible congregation of students washing hands in communal settings at school (51).

Several limitations of this study should be noted. First, the transmission patterns within schools were estimated from a single dataset of seasonal influenza in primary schools (aged 5-12 years) in Matsumoto city, Japan, and it is unclear to what extent the results can be extrapolated to other settings, e.g. secondary schools or schools in other countries. Some features of our results may still be relevant to transmission dynamics in different types of schools if they reflect general social contact behaviours of schoolchildren; however, the relative contribution of within-class/within-grade interactions may become smaller for older students (52). The data points used in the inference mostly consisted of classes of size 20-40 (those with a size smaller than 10 were excluded as they might be operated differently) and most schools had no more than 5 classes per grade. The scope of the estimated effect of the school-based interventions was also limited to within this range for internal consistency and thus may not necessarily be applicable to class structures outside this range (e.g. splitting a class of 20 students into two). Extrapolating the estimated transmission patterns to other respiratory infectious diseases also warrants caution because their epidemiological characteristics may not be identical, although we believe that such an approach may still be useful for diseases sharing similar modes of transmission. Modelling studies using social contact data often assume proportionality between contacts and the transmission of directly-transmitted diseases (e.g. measles, influenza and COVID-19) and have many successful applications (7, 33, 53–57). Using the estimated transmission patterns of influenza as a proxy for other diseases essentially rests on the same assumption, which nonetheless has limitations and should eventually be validated by disease-specific studies. Second, some aspect of the outbreaks may have been missing from the dataset. Since the illness data of teachers were not available, they were not considered throughout the analysis. However, their role in seasonal influenza transmission may have been minor given a large number of student cases and the smaller risk in adults (58, 59). Although our student incidence data likely had good case ascertainment given encouraged medical attendance and confirmation by rapid diagnostic kits (18), a certain proportion of infections (e.g. asymptomatic or very mild) may have been missing. We believe that students feeling unwell due to influenza mostly attended medical institutions and received a test as it was encouraged by schools. Nonetheless, it should be noted that this could have been a source of bias in the estimated transmission patterns. Students with very mild symptoms (e.g. only slight sore throat) may visit a medical institution only if they know of other classmates also diagnosed with influenza. If such cases were common, the contribution of within-class transmissions in our results might have been an overestimate. Third, since the dataset was obtained from an observational study, the identified determinants of transmission may not be causal and should not be viewed as conclusive evidence. The results of our log-linear regression were mostly in line with existing findings, however, our dataset may still be biased due to unmeasured confounders such as health awareness. Our estimates of the relative effect of school-based interventions were based on the assumption that students’ behaviours follow the fixed patterns according to the school structure even under interventions. That is, when the class size or the number of classes were changed by an intervention, students were assumed to change their behaviour according to the new school structure (as if it were the original structure) by e.g. rewiring close contacts in a timely manner. This is a hypothetical expectation that may not exactly be observed in actual interventional settings; for example, it may take time for students to resume close contacts after the class is split, which can bring *R*_S_ lower than our prediction at least temporarily. We have also neglected the possible effect of the interventions on the transmission outside the school. The actual effects of these interventions should ideally be validated by empirical data, as in (42).

Our analysis disentangled the transmission dynamics of seasonal influenza among primary school students and highlighted the relative importance of within-class and within-grade transmission. Since class and school sizes were minimally associated with the within-school reproduction number, school-based interventions that change classroom structures, e.g. reduced class sizes and staggered attendance, may have limited effectiveness. Empirical evidence on fine-grained heterogeneous transmission patterns at school as was obtained from this study would inform public health planning for future outbreaks of influenza and, potentially, other directly transmitted infectious diseases that thrive in schools.

## Materials and methods

### Data

We analysed a citywide school-based influenza survey data from the 2014/15 season. The survey was conducted in Matsumoto city (population size: 242,000 (60)), Japan, enrolling 13,217 students from all 29 public primary schools in the city. During the survey period (from October 2014 to February 2015), the participants were asked to fill out a questionnaire when they were back from the suspension of attendance due to diagnosed influenza (prospective survey). In March, the participants were asked to respond to another survey on their experience during the study period, regardless of whether they had contracted influenza (retrospective survey). A total of 2,548 diagnosed influenza episodes were reported in the prospective survey, which accounted for 96% of the cases officially recognised by the schools during the study period. Primary schools in Japan often requested students suspected of influenza to seek diagnosis at a medical institution. All students reporting an influenza episode in the prospective survey answered that they had received a diagnosis and at least 95% of them were noticed of type A influenza (indicating that they were lab-confirmed). In the retrospective survey, 11,390 (86%) participants responded, among which 8,375 reported that they did not have influenza during the study period.

We combined those who responded to the prospective survey (“case group”) and those who reported no influenza experience in the retrospective survey (“control group”) and obtained a dataset of 10,923 students. Of those, 71 students from 3 schools with less than 15 students per grade were excluded because they may have different schooling patterns from other schools (e.g. some students in different grades shared classrooms). We used individual profiles (sex, school, grade, class, household composition), onset dates, influenza episodes of household members and precaution measures students engaged in (vaccine, mask, hand washing) in the subsequent analysis. Further details of the dataset can be found in the original studies (51, 61).

The secondary data analysis conducted in the present study was approved by the ethics committee at the London School of Hygiene & Tropical Medicine (reference number: 14599).

### Inference model

We modelled within-school transmission considering class structures as follows. We defined the “school proximity” *d* between a pair of students *i* and *j* attending the same school as

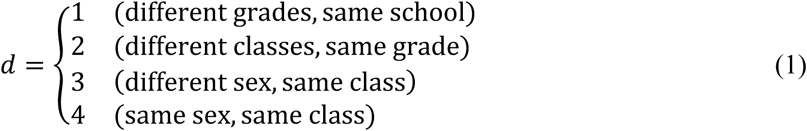

To investigate the potential effect of reduced class sizes and the number of attending students, we modelled the transmission between students as a function of two variables: the class size *n* and the number of classes per grade *m* (i.e. the number of students per grade is *nm*). Namely, we assumed that in the absence of any individual covariate effects, the cumulative transmission rate between student *i* and *j* in proximity *d* over the infectious period is represented as

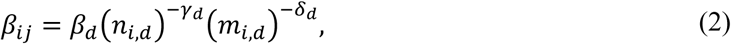

where *β*_*d*_, *γ*_*d*_, *δ*_*d*_ are parameters to be estimated. When *i* and *j* are in the same grade (i.e. *d* = 2, 3, 4), the average class size and the number of classes in that grade were used as *n*_*i,d*_ and *m*_*i,d*_. When *d* = 1, the school average was used as *n*_*i,d*_ and *m*_*i,d*_. The exponent parameters within the same class were assumed to be equal: *γ*_3_ = *γ*_4_ and *δ*_3_ = *δ*_4_.

We modelled the daily hazard of incidence for student *i* as a renewal process. Let h_τ_ be the onset-based transmission hazard as a function of serial interval *s* (normalised such that 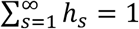; *h*_*s*_ = 0 for *s* ≤ 0). We used a gamma distribution of a mean of 1.7 and a standard deviation of 1.0 for influenza, which resulted in a mean serial interval of 2.2 days (62). The daily hazard of disease onset attributed to school transmission is given as

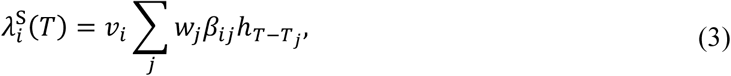

where *v*_*i*_ and *w*_*i*_ represent the relative susceptibility and infectiousness, respectively, which are specified for each individual by a log-linear regression model to account for covariates (see Supplementary materials for detailed methods).

In addition to the above within-school transmission, we also considered within-household transmission and general community transmission. Within-household transmission was incorporated as the Longini-Koopman model (63) with parameters from a previous study on the same cohort of students (18). General community transmission was modelled as a logistic curve fitted to the total incidence in the dataset to reflect the overall trend of the epidemic. See Supplementary materials for further details of the model.

We constructed the likelihood function and estimated the parameters by the Markov-chain Monte Carlo (adaptive mixture Metropolis) method. We obtained 1,000 thinned samples from 100,000 iterations after 100,000 iterations of burn-in, which yielded the effective sample size of at least 300 for each parameter. Using the posterior samples, we computed the proximity-specific reproduction number *R*_*d*_ in a hypothetical 6-year school with given *n* and *m* (assumed to be constant schoolwide) as

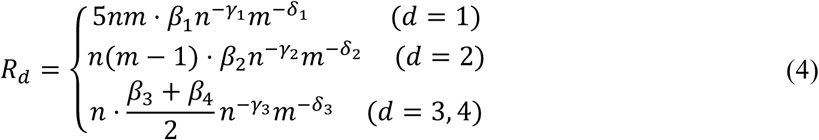

and defined the within-school reproduction number *R*_S_ as a sum of them.

We predicted the relative reduction in *R*_S_ under intervention measures changing the number of attending students and class structures by using posterior samples. Interventions were assumed to change *n* and *m* as shown in Table 1, and the predictive distribution of the relative change in *R*_S_ was computed for each intervention. The estimated *R*_S_ represents the value in a hypothetical condition where an infectious student spends the whole infectious period at school; the effect of absence due to symptoms or the staggered attendance was not included in this reduction.

All analysis was performed in Julia 1.5.2 and R 4.1.0. Replication code is available on GitHub (https://github.com/akira-endo/schooldynamics_FluMatsumoto14-15).

## Data Availability

Due to potentially sensitive information included, the original dataset is not made public and is available from the corresponding author upon reasonable request. A processed dataset with an increased level of anonymity, which can still qualitatively reproduce the main study finding (i.e. breakdown of the school reproduction number breakdown by the class/grade relationship without adjustment for covariates) is publicly available along with the accompanying code on a GitHub repository (https://github.com/akira-endo/schooldynamics_FluMatsumoto14-15).

## Acknowledgement

This research was partially funded by Lnest Grant Taisho Pharmaceutical Award. AE was financially supported by The Nakajima Foundation and The Alan Turing Institute. Yang Liu is supported by Bill & Melinda Gates Foundation [INV-003174], National Institute for Health Research [16/137/109], European Commission [101003688] and UK Medical Research Council [MC_PC_19065]. KEA is supported by European Research Council Starting Grant [757688]. AJK [206250] and SF [210758] are supported by the Wellcome Trust.

## Prior publication

Earlier version of this manuscript is available at Research Square [https://doi.org/10.21203/rs.3.rs-322366/v1].

## Footnotes

1 For example, the estimated relative reduction was 17% (95% credible interval: -16%–40%) for m = 3.

## References

1. N. A. Christakis, J. H. Fowler, Social Network Sensors for Early Detection of Contagious Outbreaks. PLOS ONE 5, e12948 (2010).

2. O. le Polain de Waroux, et al., Identifying human encounters that shape the transmission of Streptococcus pneumoniae and other acute respiratory infections (2018) https://doi.org/10.1016/j.epidem.2018.05.008.

3. S. Eubank, et al., Modelling disease outbreaks in realistic urban social networks. Nature (2004) https://doi.org/10.1038/nature02541.

4. L. A. Meyers, M. E. J. Newman, M. Martin, S. Schrag, Applying network theory to epidemics: Control measures for Mycoplasma pneumoniae outbreaks. Emerging Infectious Diseases (2003) https://doi.org/10.3201/eid0902.020188.

5. W. P. Glezen, Emerging Infections: Pandemic Influenza. Epidemiologic Reviews 18, 64–76 (1996).

6. L. Wang, et al., Transmission Characteristics of Different Students during a School Outbreak of (H1N1) pdm09 Influenza in China, 2009. Sci Rep 4, 5982 (2014).

7. K. T. D. Eames, The influence of school holiday timing on epidemic impact. Epidemiology and Infection (2014) https://doi.org/10.1017/S0950268813002884.

8. K. T. D. Eames, N. L. Tilston, E. Brooks-Pollock, W. J. Edmunds, Measured Dynamic Social Contact Patterns Explain the Spread of H1N1v Influenza. PLOS Computational Biology 8, e1002425 (2012).

9. S. Cauchemez, A.-J. Valleron, P.-Y. Boëlle, A. Flahault, N. M. Ferguson, Estimating the impact of school closure on influenza transmission from Sentinel data. Nature 452, 750–754 (2008).

10. A. J. K. Conlan, et al., Measuring social networks in british primary schools through scientific engagement. Proceedings of the Royal Society B: Biological Sciences (2011) https://doi.org/10.1098/rspb.2010.1807.

11. M. Leecaster, et al., Estimates of Social Contact in a Middle School Based on Self-Report and Wireless Sensor Data. PLOS ONE (2016) https://doi.org/10.1371/journal.pone.0153690.

12. J. Fournet, A. Barrat, Contact patterns among high school students. PLoS ONE (2014) https://doi.org/10.1371/journal.pone.0107878.

13. H. Guclu, et al., Social Contact Networks and Mixing among Students in K-12 Schools in Pittsburgh, PA. PLOS ONE 11, e0151139 (2016).

14. J. Stehlé, et al., High-resolution measurements of face-to-face contact patterns in a primary school. PLoS ONE (2011) https://doi.org/10.1371/journal.pone.0023176.

15. Simon. Cauchemez, et al., Role of social networks in shaping disease transmission during a community outbreak of 2009 H1N1 pandemic influenza. Proceedings of the National Academy of Sciences 108, 2825–2830 (2011).

16. V. Clamer, I. Dorigatti, L. Fumanelli, C. Rizzo, A. Pugliese, Estimating transmission probability in schools for the 2009 H1N1 influenza pandemic in Italy. Theoretical Biology and Medical Modelling 13, 19 (2016).

17. M. Begon, et al., A clarification of transmission terms in host-microparasite models: Numbers, densities and areas. Epidemiology and Infection (2002) https://doi.org/10.1017/S0950268802007148.

18. A. Endo, M. Uchida, A. J. Kucharski, S. Funk, Fine-scale family structure shapes influenza transmission risk in households: Insights from primary schools in Matsumoto city, 2014/15. PLoS Computational Biology (2019) https://doi.org/10.1371/journal.pcbi.1007589.

19. P. H. Thrall, A. Biere, M. K. Uyenoyama, Frequency-Dependent Disease Transmission and the Dynamics of the Silene-Ustilago Host-Pathogen System. The American Naturalist 145, 43–62 (1995).

20. E. S. Nightingale, O. J. Brady, C. C.-19 working Group, L. Yakob, The importance of saturating density dependence for predicting SARS-CoV-2 resurgence. medRxiv, 2020.08.28.20183921 (2020).

21. M. J. Smith, et al., Host–pathogen time series data in wildlife support a transmission function between density and frequency dependence. PNAS 106, 7905–7909 (2009).

22. B. Borremans, J. Reijniers, N. Hens, H. Leirs, The shape of the contact–density function matters when modelling parasite transmission in fluctuating populations. Royal Society Open Science 4, 171308.

23. N. M. Ferguson, et al., Strategies for mitigating an influenza pandemic. Nature (2006) https://doi.org/10.1038/nature04795.

24. N. M. Ferguson, et al., Strategies for containing an emerging influenza pandemic in Southeast Asia. Nature 437, 209–214 (2005).

25. L. Fumanelli, M. Ajelli, S. Merler, N. M. Ferguson, S. Cauchemez, Model-Based Comprehensive Analysis of School Closure Policies for Mitigating Influenza Epidemics and Pandemics. PLoS Comput Biol 12 (2016).

26. W. M. Getz, C. Carlson, E. Dougherty, T. C. Porco, R. Salter, An agent-based model of school closing in under-vaccinated communities during measles outbreaks. SIMULATION 95, 385–393 (2019).

27. J. Panovska-Griffiths, et al., Determining the optimal strategy for reopening schools, the impact of test and trace interventions, and the risk of occurrence of a second COVID-19 epidemic wave in the UK: a modelling study. The Lancet Child & Adolescent Health 4, 817–827 (2020).

28. M. J. Keeling, et al., The impact of school reopening on the spread of COVID-19 in England. Philosophical Transactions of the Royal Society B: Biological Sciences 376, 20200261 (2021).

29. B. Phillips, D. T. Browne, M. Anand, C. T. Bauch, Model-based projections for COVID-19 outbreak size and student-days lost to closure in Ontario childcare centres and primary schools. Sci Rep 11, 6402 (2021).

30. A. Best, et al., The impact of varying class sizes on epidemic spread in a university population. Royal Society Open Science 8, 210712.

31. A. Bilinski, J. A. Salomon, J. Giardina, A. Ciaranello, M. C. Fitzpatrick, Passing the Test: A Model-Based Analysis of Safe School-Reopening Strategies. Ann Intern Med (2021) https://doi.org/10.7326/M21-0600 (June 16, 2021).

32. S. Cauchemez, et al., Household transmission of 2009 pandemic influenza A (H1N1) virus in the United States. The New England journal of medicine (2009) https://doi.org/10.1056/NEJMoa0905498.

33. J. Mossong, et al., Social Contacts and Mixing Patterns Relevant to the Spread of Infectious Diseases. PLoS Medicine 5, e74 (2008).

34. T. Britton, T. Kypraios, P.D. O’neill, Inference for Epidemics with Three Levels of Mixing: Methodology and Application to a Measles Outbreak. Scandinavian Journal of Statistics 38, 578–599 (2011).

35. R. Christensen, Log-Linear Models and Logistic Regression, 2nd Ed. (Springer-Verlag, 1997) https://doi.org/10.1007/b97647 (June 30, 2021).

36. M. Begon, et al., A clarification of transmission terms in host-microparasite models: Numbers, densities and areas. Epidemiology and Infection (2002) https://doi.org/10.1017/S0950268802007148.

37. M. Biggerstaff, S. Cauchemez, C. Reed, M. Gambhir, L. Finelli, Estimates of the reproduction number for seasonal, pandemic, and zoonotic influenza: A systematic review of the literature. BMC Infectious Diseases (2014) https://doi.org/10.1186/1471-2334-14-480.

38. B. Killingley, J. Nguyen-Van-Tam, Routes of influenza transmission. Influenza and Other Respiratory Viruses 7, 42–51 (2013).

39. R. K. Banik, A. Ulrich, Evidence of Short-Range Aerosol Transmission of SARS-CoV-2 and Call for Universal Airborne Precautions for Anesthesiologists During the COVID-19 Pandemic. Anesthesia & Analgesia 131, e102–e104 (2020).

40. M. Klompas, M. A. Baker, C. Rhee, Airborne Transmission of SARS-CoV-2. JAMA 324, 441 (2020).

41. K. Y. Leung, F. Ball, D. Sirl, T. Britton, Individual preventive social distancing during an epidemic may have negative population-level outcomes. Journal of The Royal Society Interface 15, 20180296 (2018).

42. J. Lessler, et al., Household COVID-19 risk and in-person schooling. Science 372, 1092–1097 (2021).

43. N. Sugaya, et al., Trivalent inactivated influenza vaccine effective against influenza A(H3N2) variant viruses in children during the 2014/15 season, Japan. Eurosurveillance 21, 30377 (2016).

44. J. Howard, et al., An evidence review of face masks against COVID-19. PNAS 118 (2021).

45. B. J. Cowling, G. M. Leung, Face masks and COVID-19: don’t let perfect be the enemy of good. Eurosurveillance 25, 2001998 (2020).

46. K. Chaabna, S. Doraiswamy, R. Mamtani, S. Cheema, Facemask use in community settings to prevent respiratory infection transmission: A rapid review and meta-analysis. International Journal of Infectious Diseases 104, 198–206 (2021).

47. B. J. Cowling, Y. Zhou, D. K. M. Ip, G. M. Leung, A. E. Aiello, Face masks to prevent transmission of influenza virus: a systematic review. Epidemiology & Infection 138, 449–456 (2010).

48. H. Bundgaard, et al., Effectiveness of Adding a Mask Recommendation to Other Public Health Measures to Prevent SARS-CoV-2 Infection in Danish Mask Wearers. Ann Intern Med 174, 335–343 (2021).

49. J. Xiao, et al., Nonpharmaceutical Measures for Pandemic Influenza in Nonhealthcare Settings—Personal Protective and Environmental Measures - Volume 26, Number 5—May 2020 - Emerging Infectious Diseases journal - CDC https://doi.org/10.3201/eid2605.190994 (June 15, 2021).

50. A. E. Aiello, et al., Mask Use, Hand Hygiene, and Seasonal Influenza-Like Illness among Young Adults: A Randomized Intervention Trial. The Journal of Infectious Diseases (2010) https://doi.org/10.1086/650396.

51. M. Uchida, et al., Effectiveness of vaccination and wearing masks on seasonal influenza in Matsumoto City, Japan, in the 2014/2015 season: An observational study among all elementary schoolchildren. Preventive Medicine Reports 5, 86–91 (2017).

52. H. Guclu, et al., Social Contact Networks and Mixing among Students in K-12 Schools in Pittsburgh, PA. PLOS ONE 11, e0151139 (2016).

53. L. Munasinghe, Y. Asai, H. Nishiura, Quantifying heterogeneous contact patterns in Japan: a social contact survey. Theoretical Biology and Medical Modelling 16, 6 (2019).

54. S. Funk, et al., Combining serological and contact data to derive target immunity levels for achieving and maintaining measles elimination. BMC Medicine 17, 180 (2019).

55. D. M. Feehan, A. S. Mahmud, Quantifying population contact patterns in the United States during the COVID-19 pandemic. Nat Commun 12, 893 (2021).

56. C. I. Jarvis, et al., Quantifying the impact of physical distance measures on the transmission of COVID-19 in the UK. BMC Medicine 18, 124 (2020).

57. J. D. Munday, et al., Estimating the impact of reopening schools on the reproduction number of SARS-CoV-2 in England, using weekly contact survey data. medRxiv, 2021.03.06.21252964 (2021).

58. J. Brugger, C. L. Althaus, Transmission of and susceptibility to seasonal influenza in Switzerland from 2003 to 2015. Epidemics 30, 100373 (2020).

59. P. Arevalo, H. Q. McLean, E. A. Belongia, S. Cobey, Earliest infections predict the age distribution of seasonal influenza A cases. eLife 9, e50060 (2020).

60. Matsumoto City, Population by year (1990–2017) (June 29, 2021).

61. M. Uchida, et al., Prospective epidemiological evaluation of seasonal influenza in all elementary schoolchildren in Matsumoto city, Japan, in 2014/2015. Japanese Journal of Infectious Diseases (2017) https://doi.org/10.7883/yoken.JJID.2016.037.

62. M. A. Vink, M. C. J. Bootsma, J. Wallinga, Serial Intervals of Respiratory Infectious Diseases: A Systematic Review and Analysis. American Journal of Epidemiology 180, 865–875 (2014).

63. I. M. Longini, J. S. Koopman, Household and community transmission parameters from final distributions of infections in households. Biometrics 38, 115–126 (1982).

