## Supplementary materials for "Within and between classroom transmission patterns of seasonal influenza among primary school students in Matsumoto city, Japan"

### Supplementary materials: Within and between classroom transmission patterns of seasonal influenza and implications for pandemic management strategies at schools

Akira Endo, Mitsuo Uchida, Naoki Hayashi, Yang Liu, Katherine E. Atkins, Adam J. Kucharski, Sebastian Funk

#### Model specifications

##### Transmission model

Our dataset consisted of four components for each student: final disease outcome *D* (1 for cases and 0 for controls), onset date of students *T* (NA for controls), household data *H* (i.e. household composition and how many members have had disease) and individual covariates *X*. The likelihood for student *i* with data components $\{D_i, T_i, H_i, X_i\}$ is given as

$$\begin{cases} L_i = p(H_i|\theta^{\mathrm{H}})\Gamma_i^{\mathrm{S}}(T_{\max})\Gamma_i^{\mathrm{H}}(T_{\max})\Gamma_i^{\mathrm{C}}(T_{\max}) & (D_i = 0) \\ L_i = p(T_i, H_i|\theta^{\mathrm{H}}, \theta)\Gamma_i^{\mathrm{S}}(T_i - 1)\Gamma_i^{\mathrm{H}}(T_i - 1)\Gamma_i^{\mathrm{C}}(T_i - 1) & (D_i = 1) \end{cases} \quad \text{(S1)}$$

where $\Gamma_i^X(T)$ is the probability that student *i* survives the force of infection in settings *X* (S: school, H: household, C: general community) until time *T*. The first term of each product, $p(H_i|\theta^{\mathrm{H}})$ or $p(T_i, H_i|\theta^{\mathrm{H}}, \theta)$, represents the probability of observing household data $H_i$ and onset date $T_i$ (if $D_i = 1$) given sets of parameters $\theta^{\mathrm{H}}$ and $\theta$. The parameter set $\theta^{\mathrm{H}}$ consists of fixed parameters governing the within-household transmission, which we retrieved from the previous study on the same study cohort [1]. All other parameters $\theta$ are estimated. Since $\theta^{\mathrm{H}}$ are assumed to be fixed, the likelihoods in Equation (S1) can be simplified as

$$\begin{cases} L_i \propto \Gamma_i^{\mathrm{S}}(T_{\max})\Gamma_i^{\mathrm{C}}(T_{\max}) & (D_i = 0) \\ L_i \propto p(T_i, H_i|\theta^{\mathrm{H}}, \theta)\Gamma_i^{\mathrm{S}}(T_i - 1)\Gamma_i^{\mathrm{C}}(T_i - 1) & (D_i = 1) \end{cases} \quad \text{(S2)}$$

The survival probabilities for the school and community settings are modelled as

$$\begin{aligned} \Gamma_i^{\mathrm{S}}(T) &= \exp\left(-v_i \sum_j w_j \beta_{ij} \sum_{t=T_j+1}^{T} h_{t-T_j}\right), \\ \Gamma_i^{\mathrm{C}}(T) &= \exp(-v_i r_{\mathrm{C}} \Lambda_{\mathrm{C}}(T)), \end{aligned} \tag{S3}$$

where $\Lambda_{\mathrm{C}}(T)$ is the cumulative density function of a logistic curve representing the time trend of community outbreak (see Section "Community transmission" for details)

The likelihood for student's onset and household episodes $p(T_i, H_i|\theta^{\mathrm{H}}, \theta)$ is obtained as follows. First, the probability that student *i* has illness onset on $T_i$ due to infection either from school or general community is

$$p_i^{\mathrm{S+C}}(T_i) = 1 - \exp\left[-\left(\lambda_i^S(T_i) + \lambda^{\mathrm{C}}(T_i)\right)\right], \tag{S4}$$

where $\lambda_i^S(T_i)$ is as specified in Equation (3) in the main text and $\lambda^{\mathrm{C}}(T_i)$ represents the hazard from community outbreak given as $\lambda^{\mathrm{C}}(T) = r_{\mathrm{C}} \frac{d}{dT} \Lambda_{\mathrm{C}}(T)$. This probability $p_i^{\mathrm{S+C}}(T_i)$ is then plugged into the household transmission model. In the prospective survey, household data $H_i$ consisted of household cases simultaneously reported with the student's influenza episode. Since the illness onset dates were not reported for household cases, it was not possible to determine the direction of within-household transmissions. We assumed that the reported household cases represent those who could be linked to the student's onset (household cases infecting the student or vice versa) and thus their onset dates should be close enough to that of the student. It is also possible that they had been coprimary cases, i.e. unlinked infections separately obtained from outside the household, but we expect the onset dates to be within the same range even in such cases. Given the probability of a student acquiring disease from outside the household $p^{\mathrm{S+C}}(T_i)$, the likelihood of household data $H_i$ is given as

$$p_i(T_i, H_i) = p_i^{\mathrm{S+C}}(T_i)\pi\left((D_i = 1, H_i)\middle|\theta^{\mathrm{H}}, D_i = 1\right) + \left(1 - p_i^{\mathrm{S+C}}(T_i)\right)\pi\left((D_i = 1, H_i)\middle|\theta^{\mathrm{H}}\right), \tag{S5}$$

where $\pi$ is the likelihood of observing household final outcome $(D_i, H_i)$ (i.e. which members of the household had influenza) used in the previous study [1]. The component $\pi\left((D_i = 1, H_i)\middle|\theta^{\mathrm{H}}, D_i = 1\right)$ represents the probability of observing the household data $H_i$ given that the student is infected outside the household, and $\pi\left((D_i = 1, H_i)\middle|\theta^{\mathrm{H}}\right)$ the probability of observing $H_i$ and the student infected from the household. The household likelihood model was parameterised using the median estimates reported in [1] except the external risk of infection (see "Community transmission"). The use of median estimates to summarise the posterior samples could induce underestimation; we adopted this approximation for computational convenience, which was unlikely to have substantially affected our conclusions as the contribution of the household likelihood to the qualitative results was relatively minor (see "Additional analysis").

#### Community transmission

We fitted a logistic curve $\hat{\lambda}_C(T) = \frac{a_3}{1+\exp(-a_1(T-a_2))}$ to the aggregated incidence data of students to represent the time trend of community outbreak. The Poisson likelihood was maximised to infer the parameters $a_1$, $a_2$ and $a_3$. The fitted logistic curve was normalised, i.e. $\lambda_\mathrm{C}(T) = \frac{d}{dT}\Lambda_\mathrm{C}(T) = \frac{\hat{\lambda}_C(T)}{\int_0^\infty \hat{\lambda}_C(T)dT}$, and used in Equation S2 and as a part of household likelihood $\pi$. Since the parameter estimates for the external risk of infection (probability of infection from outside the household) in [1] corresponded to the cumulative risk over the season, we rescaled them to account for the shorter time windows in the current analysis. We assumed that influenza episodes of household members accompanying the reported episode of a student occurred within a few days range of the reported onset date of students; otherwise they might not have been reported as "coincided episodes". The external risk of infection $\varepsilon_k$ for a type $k$ household member ('sibling', 'father', 'mother', or 'other') as estimated in [1] was rescaled as

$$\varepsilon'_k = u\varepsilon_k\lambda_\mathrm{C}(T), \tag{S6}$$

where $T$ is the onset date of the student and $u$ is a constant scaling factor representing the period of exposure of household members given the onset of the student specified as a single day. We selected $u = 1$ in the main analysis because it showed a good performance in our model validation (see "Model validation by simulated data"). As a sensitivity analysis, we confirmed that different choices of $u$ ($u = 7$ and $u = 0$, which corresponds to the exclusion of household likelihood) showed overall similar patterns in the estimates of $R_d$ (see "Additional analysis").

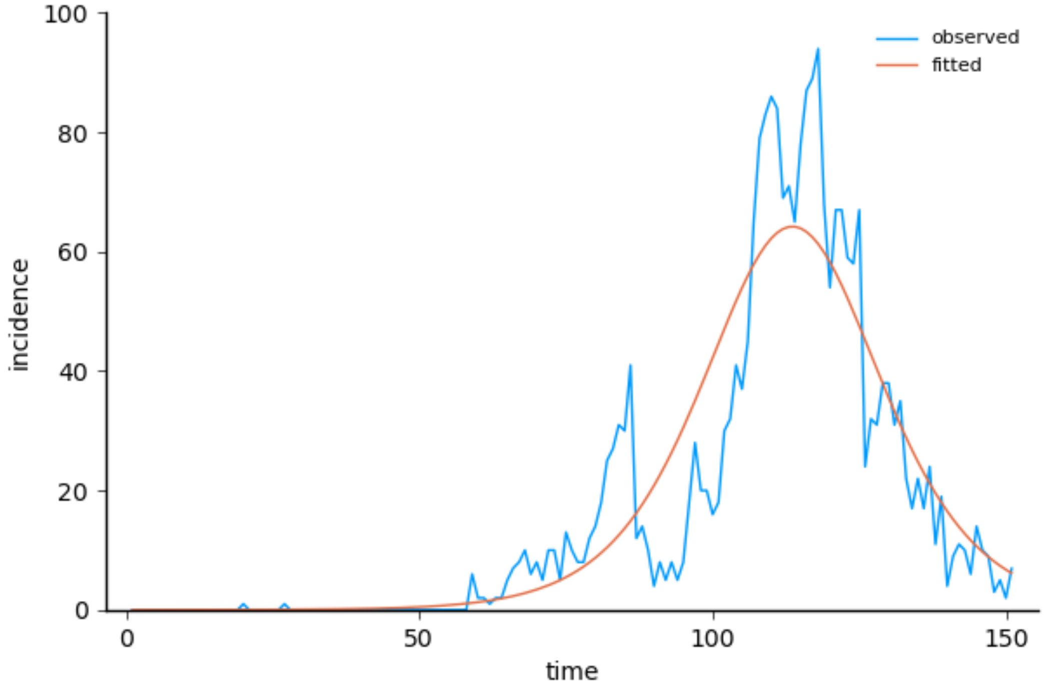


Figure S1. Logistic curve fitted to the observed incidence to model temporal trend in community transmission. The blue curve shows the aggregated incidence of students. The x-axis represents dates of illness onset, where day 1 corresponds to 1 October 2014.

#### Adjustment for covariates

Covariates $\boldsymbol{\xi}_i = \left(\xi_i^1, \xi_i^2, \ldots\right)$ that may affect the susceptibility or infectiousness (shown in Table S2) were addressed by log-linear regression:

$$
\begin{aligned}
v_i &= \exp\left(\boldsymbol{\xi}_i^{\mathrm{T}} \boldsymbol{\alpha}_i^v\right), \\
w_i &= \exp\left(\boldsymbol{\xi}_i^{\mathrm{T}} \boldsymbol{\alpha}_i^w\right),
\end{aligned}
\tag{S7}
$$

where $\boldsymbol{\alpha}_i^v$ and $\boldsymbol{\alpha}_i^w$ denote vectors of regression coefficients. We assumed that the variables $v_i$ and $w_i$ are involved in school and community transmission and not in household transmissions for methodological convenience, based on our assumption that protective effects including precaution measures may not be as effective inside households as outside. We believe this assumption is unlikely to have biased our inference on within and between class transmission patterns because exclusion of the household likelihood barely changed the estimates in our sensitivity analysis (see "Additional analysis").

Since the covariates included self-reported precaution measures (vaccination, mask and hand washing), there were mismatches between the responses in the prospective and retrospective surveys that warranted our attention. Due to the different nature of recruitment (prospective survey accompanied leave-of-absence forms while retrospective survey was a mass questionnaire), the retrospective survey had a lower respondent rate (86%) than the prospective survey (96%). Limiting to those reporting influenza episodes ('cases') who were eligible to both the prospective and retrospective surveys, the retention rate (the proportion of prospective survey respondents who remained in the retrospective survey) was 84% overall. However, the retention rate by responses on the precaution measures showed inconsistent patterns (Table S1); more positive responses remained in the prospective surveys on "vaccination" and "mask" and vice versa on "hand washing". Notably, the retention rate of "hand washing: No" is over 100%, suggesting that there were students who answered "No" only in the retrospective survey (as the records were unlinkable between the surveys, it was not possible to confirm which students had mismatched responses). Although this might be explained by differential retention rates (i.e. those answering "Yes" on vaccination and masks and those answering "No" on hand washing were more likely to remain in the retrospective survey), more plausible would be that in the retrospective survey, students can give answers different from the prospective survey. This may be due to the recall bias because the retrospective survey was conducted in March, which could have been up to 4-5 months after the students' onset, and can cause biased estimates because the responses of the control group, only available for the retrospective survey, may also be inconsistent with their actual behaviour during the outbreak. We assumed that the responses of the case group in the prospective survey reflected the actual behaviours of students during the epidemic season and that the answers of both case and control groups in the retrospective survey are potentially misclassified. By treating the responses in the prospective survey as a reference, the sensitivity and specificity of responses in the retrospective survey can be estimated. Assuming nondifferential misclassification with shared sensitivity and specificity across variables, we assessed and adjusted for recall bias in the dataset.

The sensitivity and specificity of the responses in the retrospective survey was jointly estimated with the retention rates in a form of a matrix $M$ representing probabilities of retention and classification. Let $N^{\mathrm{p}}_{\xi=x}$ and $N^{\mathrm{r}}_{\xi=x}$ be the number of responses $x$ ("Yes": $x$ = 1; "No": 0) on a covariate $\xi$ in the prospective and retrospective surveys, respectively. The expectancy of $N^{\mathrm{r}}_{\xi=x}$ can be given as

$$\begin{pmatrix} E\left(N^{\mathrm{r}}_{\xi=0}\right) \\ E\left(N^{\mathrm{r}}_{\xi=1}\right) \end{pmatrix} = M \begin{pmatrix} N^{\mathrm{p}}_{\xi=0} \\ N^{\mathrm{p}}_{\xi=1} \end{pmatrix}, \quad \text{(S8)}$$

and we maximised the corresponding Poisson likelihood to estimate $M$. The responses in the prospective survey reconstructed from those in the retrospective survey and the inverted matrix $M^{-1}$ were overall consistent with the observed responses (Table S1).

Table S1. Comparison of observed and reconstructed covariates

| Covariate | Survey responses | | | Model prediction |
|---|---|---|---|---|
| | Prospective | Retrospective | Retention rate | Prospective (reconstructed) |
| Vaccination | Yes: 1122 | Yes: 978 | 87% | Yes: 1102 |
| | No: 1426 | No: 1171 | 82% | No: 1446 |
| Mask | Yes: 1204 | Yes: 1069 | 89% | Yes: 1226 |
| | No: 1344 | No: 1080 | 80% | No: 1322 |
| Hand | Yes: 2200 | Yes: 1778 | 81% | Yes: 2199 |
| washing | No: 348 | No: 371 | 107% | No: 349 |
| Total | 2548 cases | 2149 cases | 84% | — |

We used the estimated parameter matrix $M$ to adjust the likelihood function of the control group, whose responses are, by definition, missing in the prospective survey. We used the covariates of the case group as reported in the prospective survey and thus adjustment was not necessary for them. The component of the likelihood which can be affected by this adjustment is given as $\Gamma_i^{\mathrm{S}}(T_{\max})\,\Gamma_i^{\mathrm{C}}(T_{\max})$ as in Equation (S3). Noting that only $v_i$ is relevant to the adjustment, we get

$$\Gamma_i^{\mathrm{S}}(T_{\max})\,\Gamma_i^{\mathrm{C}}(T_{\max}) = \exp\left(-v_i\Lambda_i^{\mathrm{Total}}\right) = \exp\left(-\Lambda_i^{Total}\,exp\left(\boldsymbol{\xi}_i^T\boldsymbol{\alpha}_i^v\right)\right), \qquad \text{(S9)}$$

where $\Lambda_i^{\mathrm{Total}} = r_{\mathrm{C}}\Lambda_{\mathrm{C}}(T_{\max}) + \sum_j w_j\beta_{ij}\sum_{t=T_j+1}^{T_{\max}} h_{t-T_j}$ (hereafter, let us limit $\boldsymbol{\xi}$ to the three covariates shown in Table S1, where the value of 1/0 indicates Yes/No, respectively).

We then accounted for potential misclassification in the recorded covariates that determined $v_i$ by incorporating an adapted version of the multiple overimputation method [2]. For each covariate, we estimated the Bayesian probabilities $p\left(\boldsymbol{\xi}_i\middle|\hat{\boldsymbol{\xi}}_i\right)$, the conditional probability of the true values of binomial variables given the data. Although variables are repeatedly imputed in the original multiple overimputation method, we instead directly obtained the (approximated) adjusted likelihood for computational convenience as

$$\begin{aligned}
\Gamma_i^{\mathrm{S}}(T_{\max})\,\Gamma_i^{\mathrm{C}}(T_{\max}) &= \sum_{\xi_i} p(\boldsymbol{\xi}_i|\hat{\boldsymbol{\xi}}_i)\exp\left(-\Lambda_i^{Total}\,exp\left(\boldsymbol{\xi}_i^T\boldsymbol{\alpha}_i^v\right)\right) \\
&\approx \exp\left(-\Lambda_i^{Total}\sum_{\xi_i} p\left(\boldsymbol{\xi}_i\middle|\hat{\boldsymbol{\xi}}_i\right)exp\left(\boldsymbol{\xi}_i^T\boldsymbol{\alpha}_i^v\right)\right) \qquad \text{(S10)} \\
&= \exp\left(-\Lambda_i^{Total}\prod_k\left[p\left(\xi_i^k=0\middle|\hat{\xi}_i^k\right)+p\left(\xi_i^k=1\middle|\hat{\xi}_i^k\right)exp\left(\alpha_i^{v,k}\right)\right]\right).
\end{aligned}$$

We used the property of approximate linearity assuming $\left|\Lambda_i^{Total}\,exp\left(\boldsymbol{\xi}_i^T\boldsymbol{\alpha}_i^v\right)\right| \ll 1$ and also assumed an independence in misclassification.

#### Addressing sampling bias between case and control groups

Due to the lower respondent rate in the retrospective survey, the original likelihood directly constructed from the raw data underrepresented the control group. Although individual-level data (e.g. covariates and household episodes) was not available for students missing in the control group, it was still possible to estimate the number of such students as both the class sizes and the number of cases in each class were known. To avoid the overestimation of transmission risks this sampling bias could cause, we rescaled the likelihood of the control groups assuming that the individual-level data of included students are also representative of those missing. We did not consider students missing in the case group as the response rate was sufficiently high in the prospective survey (> 95%).

The adjusted likelihood of students in a class $A$ of size $n_A$, where $x_A$ cases and $y_A$ controls are observed is given as

$$\prod_{i \in A} L'_i = \left( \prod_{i \in A, D_i = 0} L_i^{\frac{n_A - x_A}{y_A}} \right) \left( \prod_{i \in A, D_i = 1} L_i \right). \tag{S11}$$

The first product represents the likelihood of the control group in the class and the second product represents that of the case group. Although this can lead to overconfidence in the log-linear regression results, the degree of such effect is minimal (e.g. 15% inflation of samples only cause 5% underestimation of standard error) and we prioritised reducing bias in transmission risk estimates.

### Additional analysis

#### Parameter lists and credible intervals

Parameter estimates and their 95% credible intervals for the main analysis are listed in Table S2 (also see Table 2 in the main text for the estimated coefficients for the log-linear regression). Note that for the sake of interpretability, the estimates for the cumulative transmission rates $\beta_d$ were rescaled such that they correspond to the rates in a school with 3 classes per grade of 30 students ($n$ = 30, $m$ =3). The improper flat priors were assumed for each parameter (or their logarithm).

Table S2. Parameter estimates

| Parameter | Notation | Transform | Median estimate (95% CrI) |
|---|---|---|---|
| Cumulative transmission rate (rescaled) | $\beta_1$ | Log | 0.00040 (0.00029–0.00055) |
| | $\beta_2$ | | 0.0013 (0.0008–0.0019) |
| | $\beta_3$ | | 0.016 (0.012–0.021) |
| | $\beta_4$ | | 0.018 (0.014–0.025) |
| Exponent for class size | $\gamma_1$ | — | 1.7 (0.2–3.3) |
| | $\gamma_2$ | | 0.9 (-1.6–3.1) |
| | $\gamma_3$ | | 1.1 (0.6–1.6) |
| Exponent for the number of classes per grade | $\delta_1$ | — | 1.0 (0.6–1.4) |
| | $\delta_2$ | | -0.01 (-0.84–0.74) |
| | $\delta_3$ | | 0.17 (0.01–0.33) |
| Risk from community | $r_C$ | Log | 0.022 (0.017–0.027) |

Log: parameters are log-transformed when supplied to the model.

#### Model comparison by Laplace-approximated model evidence

In our main analysis, we assumed that the within-school transmission rate is modified by the effect of class size and the number of classes per grade to the power of exponent parameters $\gamma$ and $\delta$, respectively; i.e. $\beta_{ij} = \beta_d \left(n_{i,d}\right)^{-\gamma_d} \left(m_{i,d}\right)^{-\delta_d}$. This "full model" assumes that the value of the exponents varies between different school proximity level $d$ and thus contains six free parameters for the exponents ($\gamma_1$, $\gamma_2$, $\gamma_3$, $\delta_1$, $\delta_2$, $\delta_3$). However, this may have led to overfitting due to the high model flexibility and simpler alternative models may exhibit better fit to the data. To address this issue, we compared models with different complexity with regards to the within-school transmission and selected the best model based on the Bayesian model evidence [3].

The Bayesian model evidence is a consistent criterion for model selection. However, the exact Bayesian model evidence is often analytically and computationally intractable and approximation methods need to be employed. The Bayesian information criterion (BIC) provides an approximation of the Bayesian model evidence if the samples are independent and identically distributed and the model is regular [4]. However, due to the interdependent nature of our dataset (i.e. infection events among students), BIC is unlikely to provide a reliable approximation of the model evidence. We instead directly approximated the model evidence using the Laplace approximation (matched to the scale of BIC) [3]:

$$E = -2\log\phi(\theta_0) - 2\log L(\theta_0) - K\log 2\pi + \log|F|, \tag{S12}$$

where $\theta_0$ is the maximum a posteriori (MAP) estimate, $\phi(\theta_0)$ and $L(\theta_0)$ are the prior density and the total likelihood at the MAP estimate, respectively, $K$ is the parameter dimension and $|F|$ is the determinant of the Fisher information matrix $F$. In our model, we used an improper flat prior $\phi(\theta) = 1$; therefore the first term of Equation (S12) is 0. The quantile-quantile (Q-Q) plot of the MCMC samples produced by the full model suggested that the posterior distribution is well approximated by a multivariate normal distribution (Figure S2), which supports the use of the Laplace approximation.

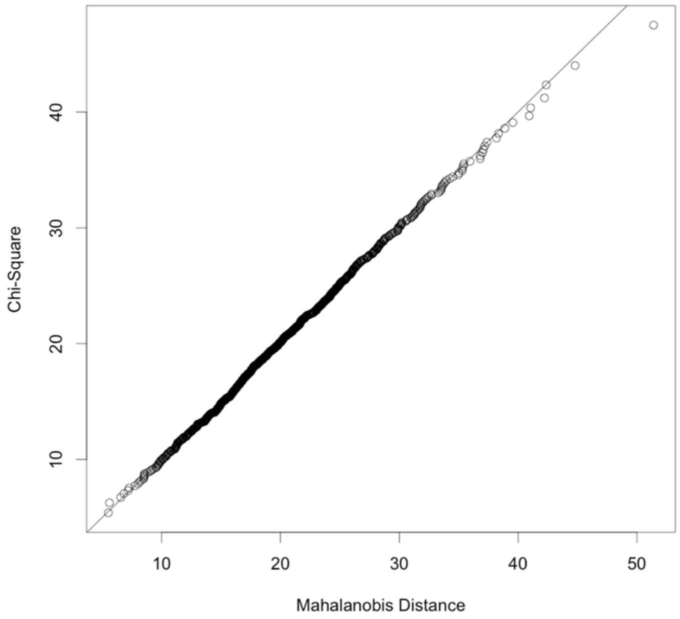


Figure S2. Quantile-quantile (Q-Q) plot for assessment of multivariate normality of the posterior samples. The sample Mahalanobis squared distances are plotted against the Chi-square quantiles (dots). The solid line represents the expected relationship for the multivariate normal distribution.

We compared our full model and its seven nested models using the Laplace-approximated model evidence (LAME) as defined in Equation (S12) (Table S3). Each of the nested models was defined by imposing certain parameter constraints to the full model and thus has fewer free parameters. None of the models considered had the least eigenvalue of the Fisher information matrix that is close to zero, i.e. the Fisher information matrix is non-degenerate, which is the necessary condition for the use of LAME. LAME strongly supported our full model (Model 0), with differences no less than 4 [5] from other models.

Table S3. Model comparison based on Laplace-approximated model evidence.

| Model ID | Parameter constraints | $K$ | $-\log L(\theta_0)$ | $\lambda_{min}$ | LAME | $\Delta$LAME |
|---|---|---|---|---|---|---|
| Model 0 | None (“full model”) | 20 | 14563.5 | 0.7 | 29173.2 | 0 (best model) |
| Model 1 | $\gamma_1 = \gamma_2 = \gamma_3$ | 18 | 14563.8 | 5.8 | 29177.5 | 4.3 |
| Model 2 | $\delta_1 = \delta_2 = \delta_3$ | 18 | 14569.5 | 0.8 | 29184.0 | 10.8 |
| Model 3 | $\gamma_1 = \gamma_2 = \gamma_3$, $\delta_1 = \delta_2 = \delta_3$ | 16 | 14571.7 | 9.8 | 29192.1 | 18.9 |
| Model 4 | $\gamma_1 = \gamma_2 = \gamma_3 = 0$, $\delta_1 = \delta_2 = \delta_3 = 0$ | 14 | 14611.2 | 10.1 | 29266.0 | 92.8 |
| Model 5 | $\gamma_1 = \gamma_2 = \gamma_3 = 1$, $\delta_1 = \delta_2 = \delta_3 = 1$ | 14 | 14634.7 | 8.5 | 29312.8 | 139.6 |
| Model 6 | $\gamma_1 = \gamma_2 = \gamma_3 = 0$, $\delta_1 = \delta_2 = \delta_3 = 1$ | 14 | 14633.2 | 8.4 | 29309.8 | 136.6 |
| Model 7 | $\gamma_1 = \gamma_2 = \gamma_3 = 1$, $\delta_1 = \delta_2 = \delta_3 = 0$ | 14 | 14588.1 | 10.2 | 29219.9 | 46.4 |

$K$: parameter dimension; $-\log L(\theta_0)$: negated maximum log likelihood; $\lambda_{min}$: the least eigenvalue of the Fisher information matrix; LAME: Laplace-approximated model evidence; ΔLAME: difference in LAME from the best model.

#### Sensitivity analysis of within and between class transmission patterns

We assessed the robustness of the estimated transmission patterns of seasonal influenza within and between classes to variations in the following assumptions: (i) serial interval distribution (ii) control of covariates (iii) use of household transmission model (iv) window period length for the household model. For each of these, we performed sensitivity analysis as described below and compared the results of estimated transmission patterns.

(i) Serial interval distribution: We used the mean serial interval of 2.2 days as estimated in [5], in the main analysis, which is slightly shorter than other estimates [6, 7]. We instead used a longer serial interval (3.5 days) as a sensitivity analysis.

(ii) Control of covariates: The log-linear regression to adjust for covariates was excluded.

(iii) Use of household transmission model: The likelihood compartment accounting for household transmission was excluded from analysis.

(iv) Scaling factor $u$ = 7 for the household model: We assumed a scaling factor $u$, which is used to weight the contribution of the risk of infection for household members from outside the household to the household likelihood. We used a larger value ($u$ = 7) than in the main analysis ($u$ = 1).

Overall, our sensitivity analysis suggested that these assumptions had limited effects on the qualitative interpretation of our results on the within and between class transmission patterns of seasonal influenza (Figure S3).

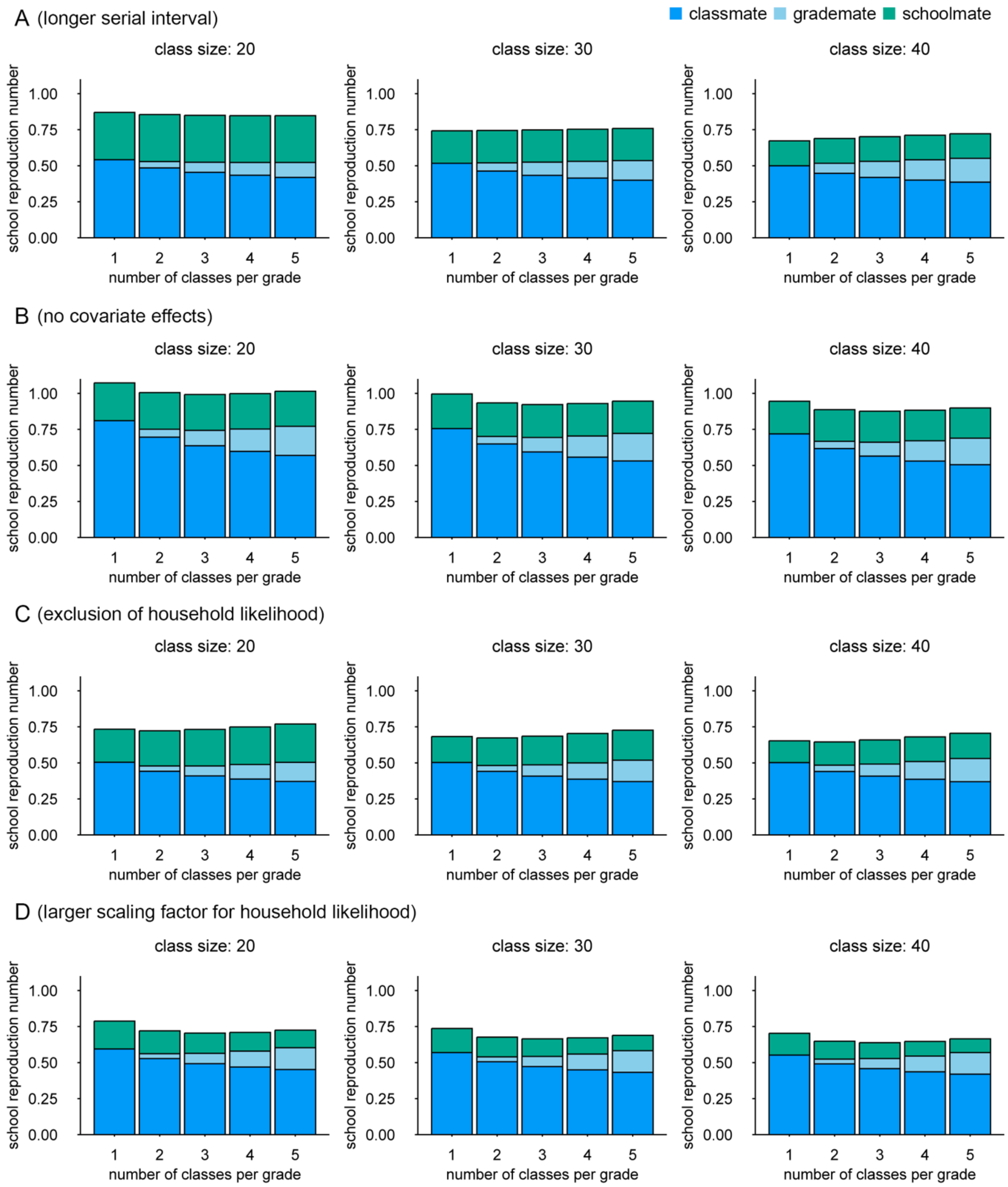


Figure S3. School reproduction number ($R_S$) and its breakdown by the class/grade relationship (median estimates) corresponding to various settings in the sensitivity analysis. (A) A longer serial interval (mean 3.5 days) was used instead of a mean 2.2 days used in the main analysis. (B) The log-linear regression used to adjust for covariates was excluded from analysis. (C) Household transmission model was excluded from the likelihood ($u = 0$). (D) A larger scaling factor was used to weight the contribution of the external infection risk for household members to the household likelihood ($u = 7$ days instead of $u = 1$ in the main analysis).

#### Sensitivity analysis excluding the recall bias adjustment

In addition to the main analysis using the recall bias adjustment (as described in "Adjustment for covariates"), we also performed the log-linear regression without the adjustment as a sensitivity analysis. The results were overall similar to the original results (Table S4), although the negative effect of hand washing on the susceptibility was slightly exaggerated.

Table S4. Log-linear regression results without the recall bias adjustment

| Covariate | Frequency in data | Relative susceptibility | Relative infectiousness |
|---|---|---|---|
| Grade (1 year) | — | 1.12 (0.91–1.40) | 0.78 (0.59–1.06) |
| Vaccine | 47.7% | 0.81* (0.75–0.89) | 0.98 (0.82–1.17) |
| Mask wearing | 51.4% | 0.70* (0.64–0.77) | 0.67* (0.57–0.79) |
| Hand washing | 80.1% | 1.95* (1.72–2.24) | 1.24 (0.95–1.67) |
| Onset in winter break | 5.9% (of cases) | — | 0.25* (0.15–0.39) |

Values are median estimates and 95% credible intervals.

* Estimates with 95% credible intervals not crossing 1.

#### Model validation by simulated data

We used a simple school-household-community outbreak simulation model to validate the performance of our inference approach that combines the likelihoods of students with onset dates and those of household members without. An outbreak was simulated among a class of students each of whom was assumed to have four other household members. Transmission was assumed to occur only between students of the same class or between members of the same household. All individuals (students and their household members) were also at the risk of infection from the general community. The serial interval distribution and the temporal distribution of the community risk were identical to those used in the main analysis (i.e. mean serial interval of 2.2 days and a logistic curve shown in Figure S1). The within-household reproduction number was fixed at 0.1, irrespective of the combination of the transmission pair. Within-school (class) transmission was specified as $\beta_{ij} =$

$\beta_3 n_3^{-\gamma_3}$, where $n_3$ is the class size. The cumulative risk of infection from the community was $r_C$ for all individuals.

Outbreaks were simulated independently in 300 classes (100 classes of size 20, 100 classes of size 30 and 100 classes of size 40) to produce one set of data for inference. The onset dates of household members were discarded and only the number of cases in the household was used for inference as in the main analysis. The parameters of interest, i.e. within-school transmissibility constant $\beta_3$, exponent parameter $\gamma_3$ and the cumulative risk of community infection $r_C$ were set at 0.05, 0.5 and 0.02, respectively and the inference model in Equation (S2) (where the school transmission is limited to within-class) was fitted to the data to estimate these parameters. In the household likelihood, the reproduction number of 0.1 and the external risk of infection 0.02 were assumed to be known; however, the scaling factor $u$ for community risk of infection ( $\varepsilon_k$ in Equation (S6)) was varied ($u$ = 1, 0 or 7) for comparison. This process of simulation and inference was repeated 50 times and the maximum likelihood estimates and the 95% confidence interval derived from the Fisher matrix were compared against the true values.

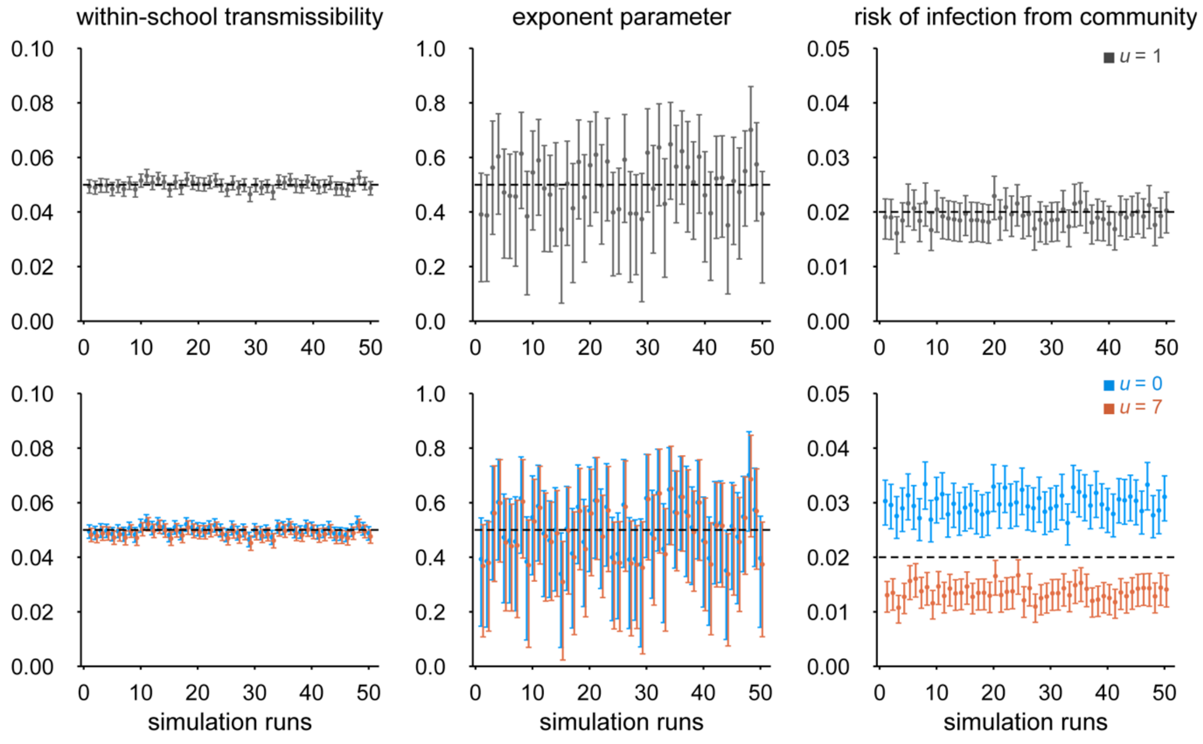

Figure S4. Model validation results using simulated data. The dots and whiskers represent the maximum likelihood estimate and the 95% confidence interval from the 50 simulations. The dashed horizontal lines denote the true parameter values used in the simulation. Top panels: parameter estimates with the scaling factor $u$ =1 (default) for the household likelihood. Bottom panels: parameter estimates with the scaling factor $u$ =0 (exclusion of household likelihood) and $u$ = 7 (overestimated risk of infection from outside the household for household members).

The simulation results suggested that our model plausibly recovered the true parameter values when $u$ = 1 (Figure S4). Although different values of $u$ ($u$ = 0 and $u$ = 7) produced slightly biased estimates for the cumulative risk of infection from the community ($r_C$), bias in the other parameter estimates for the school transmission remained minimal. We also tested a different set of parameter values: ($\beta_3$, $\gamma_3$, $r_C$) = (0.08, 0.8, 0.04) and obtained consistent results. These outcomes suggest that the use of $u$ = 1 is supported for our model and that the estimates of the parameters for the school transmission may be robust even if the household likelihood is slightly misspecified (Figure S5).

The interpretation of the best performing assumption of $u$ = 1 may not be straightforward as this essentially assumes that the household members included in the data are exposed to the risk of infection from outside the household (external risk of infection) for only one day (or the equivalent amount), although their onset dates are not specified in the likelihood (and thus can range a few days around the onset date of the student). Here we discuss a possible explanation for this. Let us consider a hypothetical scenario where the serial interval is always exactly three days; i.e. if the onset date of a student is $t$, a household member has to have an onset date of $t – 3$ to have (directly) infected the student. Since any other onset date for a household member excludes the possibility of transmission from the household member to the student, the external risk of infection for only one day is relevant. While this is not true for household members who do not directly transmit to the student (e.g. those indirectly involved in the transmission to the student or those infected by the student), the relative contribution of the external risk of infection for these household members to the likelihood is likely to be minor. As a result, $u$ = 1 may have provided a sufficient approximation of the household likelihood. The same applies to the case of the general serial interval as long as the daily risk of external infection is almost constant within the possible range of the serial interval.

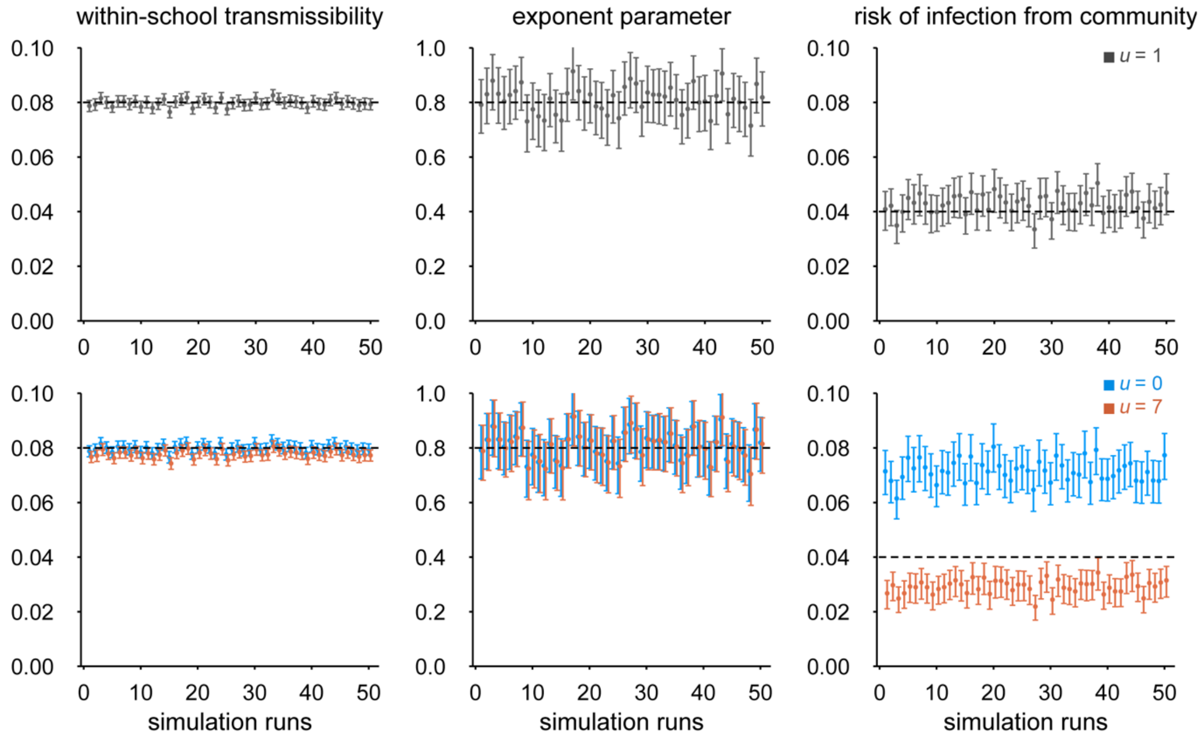


Figure S5. Model validation results using simulated data with a different set of true parameter values. The dots and whiskers represent the maximum likelihood estimate and the 95% confidence interval from the 50 simulations. The dashed horizontal lines denote the true parameter values used in the simulation. Top panels: parameter estimates with the scaling factor $u$ =1 (default) for the household likelihood. Bottom panels: parameter estimates with the scaling factor $u$ =0 (exclusion of household likelihood) and $u$ = 7 (overestimated risk of infection from outside the household for household members).

#### Reproducing temporal clustering patterns of onset dates by simulation

We assessed whether our within-school transmission model can reproduce the temporal clustering patterns of onset dates of students observed in our data (Figure 1D in the main text). We extracted 50 sets of posterior samples from the MCMC chain and simulated outbreaks among students of 26 primary schools (i.e. schools included in the main analysis) replicated from the Matsumoto city dataset over the duration of the study period (from 1 October 2014 to 28 February 2015). The risk of infection from the community and within-school transmission at four levels (see Equation (1) in the main text) were considered. The simulated onset dates were then partitioned by class, grade, school and overall and the distribution of the deviation from the group mean in 50 simulation runs are displayed in Figure S6. The simulated onset dates of students showed similar patterns to the observed distributions and the standard deviations are overall in line (Table S5).

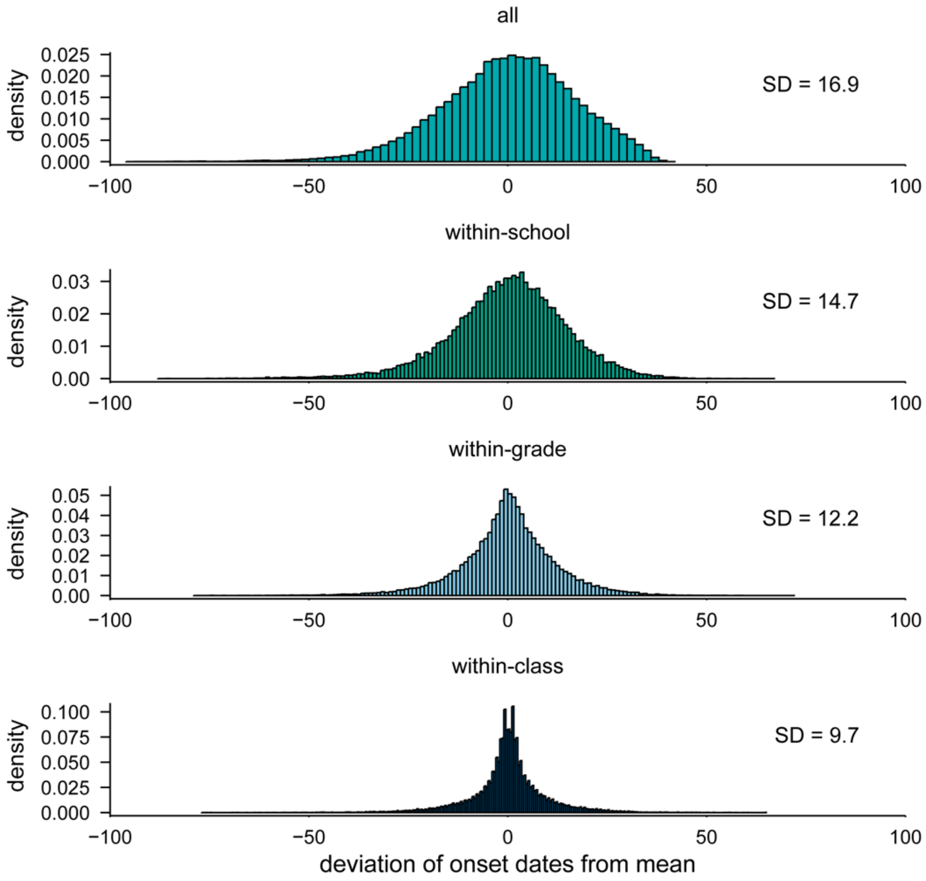


Figure S6. Temporal clustering patterns of students' onset dates with different levels of groupings reproduced from the school transmission model. The distributions of the deviance of each student's onset from the group mean are displayed at overall, school, grade and class levels. The standard deviation (SD) of each distribution is also shown.

Table S5. Comparison of observed and simulated by-group standard deviations.

| | Overall | Within-school | Within-grade | Within-class |
|---|---|---|---|---|
| Observed SD | 17.4 | 14.4 | 12.4 | 10.6 |
| Simulated SD | 16.9 | 14.7 | 12.2 | 9.7 |

SD: standard deviation of the distribution

#### References


1. Endo A, Uchida M, Kucharski AJ, Funk S. Fine-scale family structure shapes influenza transmission risk in households: Insights from primary schools in Matsumoto city, 2014/15. PLoS Comput Biol. 2019.

2. Blackwell M, Honaker J, King G. A Unified Approach to Measurement Error and Missing Data: Overview and Applications. Sociol Methods Res. 2017.

3. Knuth KH, Habeck M, Malakar NK, Mubeen AM, Placek B. Bayesian evidence and model selection. Digit Signal Process. 2015;47:50–67.

4. Schwarz G. Estimating the Dimension of a Model. Ann Stat. 1978;6. doi:10.1214/aos/1176344136.

5. Burnham KP, Anderson DR. Multimodel Inference: Understanding AIC and BIC in Model Selection. Sociol Methods Res. 2004;33:261–304.

6. Vink MA, Bootsma MCJ, Wallinga J. Serial Intervals of Respiratory Infectious Diseases: A Systematic Review and Analysis. Am J Epidemiol. 2014;180:865–75.

7. Cowling BJ, Fang VJ, Riley S, Malik Peiris JS, Leung GM. Estimation of the serial interval of influenza. Epidemiology. 2009.

8. Levy JW, Cowling BJ, Simmerman JM, Olsen SJ, Fang VJ, Suntarattiwong P, et al. The serial intervals of seasonal and pandemic influenza viruses in households in Bangkok, Thailand. Am J Epidemiol. 2013.
